# Prospective investigation of SARS-CoV-2 seroprevalence in relation to natural infection and vaccination between October 2020 and September 2021 in the Czech Republic

**DOI:** 10.1101/2022.07.21.22277881

**Authors:** Vojtěch Thon, Pavel Piler, Tomáš Pavlík, Lenka Andrýsková, Kamil Doležel, David Kostka, Hynek Pikhart, Martin Bobák, Jana Klánová

**Affiliations:** RECETOX, Faculty of Science, Masaryk University, Brno, Czech Republic; Institute of Biostatistics and Analyses, Faculty of Medicine, Masaryk University, Brno, Czech Republic; QualityLab Association, Prague, Czech Republic; Health Insurance Company of the Ministry of the Interior of the Czech Republic, Prague, Czech Republic; Department of Epidemiology & Public Health, University College London, London WC1E 6BT, UK

**Author notes:** **Corresponding author:** Prof. Vojtěch Thon, M.D., Ph.D., RECETOX, Faculty of Science, Masaryk University, Brno, Czech Republic. These authors contributed equally (Vojtěch Thon, Pavel Piler).

**Keywords:** SARS-CoV-2, seroprevalence, vaccination, epidemic growth, antibodies durability

## Abstract

**Purpose:** Monitoring the seroprevalence of COVID-19 antibodies is an important tool to design and adjust preventive strategies. We used prospective data on a large population sample with repeated IgG antibody measurement to examine changes in seropositivity before and during the national vaccination campaign in the Czech Republic.

**Methods:** 22,130 persons provided blood samples for COVID-19 IgG antibody measurement at two time points approximately 5-7 months apart, between October 2020 and March 2021 (Phase 1, before vaccination), and between April and September 2021 (during vaccination campaign).

**Results:** Before vaccination (Phase 1), seroprevalence increased from 15% in October 2020 to 56% in March 2021. By the end of Phase 2, in September 2021, prevalence increased to 91%; the highest seroprevalence was seen among vaccinated persons with and without previous SARS-CoV-2 infection (99.7% and 97.2%, respectively), while the lowest seroprevalence was found among unvaccinated persons with no signs of disease (26%). Vaccination rates were lower in persons who were seropositive in phase 1 but increased with age and body mass index. Only 9% of unvaccinated subjects who were seropositive in phase 1 became seronegative by phase 2.

**Conclusion:** The rapid increase in seropositivity during the 2^nd^ wave of the COVID-19 epidemic (covered by phase 1 of this study) was followed by a similarly steep rise in seroprevalence during the national vaccination campaign, reaching seropositivity rates of over 97% among vaccinated persons.

## Introduction

During the COVID-19 pandemics, monitoring of the seroprevalence of antibodies in the population is an important tool to design and adjust preventive strategies. As a part of this process, it is essential to assess the contribution of natural infections and vaccination to the immune response to SARS-CoV-2. The Serotracker platform has recorded hundreds of SARS-CoV-2 serological studies worldwide (serotracker.com)[1]. Most national seroprevalence studies were performed before the start of massive vaccination programme in Europe[2] but there are only few published European seroprevalence studies covering both pre- and after vaccination campaign periods. Overall, these studies, mainly based in Western Europe, reported rising seroprevalence after the national vaccination programmes[3-7]. However, very few published studies have been conducted in Central and Eastern Europe, where the dynamics of both the epidemics and vaccine uptake differed from the Western European countries.

We have previously reported findings from a national cross-sectional survey of 30,000 persons in the Czech Republic who were examined between October 2020 and March 2021, a period covering the second wave of the epidemic, which was also the period before the start of national vaccination campaign. We found that by March 2021, 53% of participants had measurable antibodies against SARS-CoV-2[8]. This was consistent with governmental data using cumulative PCR testing data. These rates were considerably higher than those reported in Western Europe[2 9-11], due to a strong 2^nd^ wave of natural infection in the Czech Republic in autumn 2020[8].

In this report, we report longitudinal data on repeated assessment of the same population sample in the period April 2021-Sept 2021, a period coinciding with the rollout of the national vaccination programme. The objectives of this analysis were to 1) examine the trends in seropositivity before and during the national vaccination campaign, 2) assess the contributions of natural infections and vaccination to the seropositivity, 3) to assess seroconversion rates in previously seronegative persons, 4) to assess duration of immunity after natural infection, and 5) to estimate the rate ratio of seroconversion and vaccination associated with sociodemographic indicators.

## Methods

### Study design and participants

Data for these analyses were derived from the first and second wave of the PROSECO study. The PROSECO study design and population recruitment has been described elsewhere[8]. Briefly, phase 1 of the study recruited 30,054 unvaccinated adult volunteers from persons registered with the second largest health insurance company in the Czech Republic. Participants provided blood sample between October 2020 and March 2021, during the 2^nd^ epidemic wave in the Czech Republic. Of those, 22,130 participants were re-examined during the national vaccination programme between April 2021 and September 2021. Participants were invited for phase 2 in the same order as they participated in phase 1, so most subjects were re-examined 5-7 months after the first visit. Comparison of the persons participating in both phases with those who only attended phase 1 is shown in **Supplementary Table S1**. Those who participated in both assessments were older, more likely to be female, seropositive at phase 1, more obese, and more likely to have history of chronic non-communicable diseases.

In phase 2, participants provided a second blood sample for detection of IgG antibodies against SARS-CoV-2 and completed a questionnaire on personal information, self-reported results of RT-PCR tests (if performed) and records of COVID vaccination. The second visit was organised at least 14 days after any vaccination (if completed). Informed consent forms were obtained from all study participants during each wave of the data collection. The study, including all aspects of data collection and data analysis, was approved by the ELSPAC ethics committee under reference number (C)ELSPAC/EK/5/2021.

### Laboratory analyses

CE-marked serological tests were performed in accredited clinical laboratories. Antigen-specific humoral immune response was analysed by detection of IgG antibodies against the spike protein using commercial immunoassays LIAISON SARS-CoV-2 S1/S2 IgG (DiaSorin, Saluggia, Italy) and SARS-CoV-2 IgG II Quant (Abbott, Sligo, Ireland). Testing was conducted on the LIAISON XL (DiaSorin, Saluggia, Italy) and on the Alinity (Abbott, Lake Forest, IL, USA) respectively. Samples were tested individually and reported according to the manufactures’ criteria.

### Statistical analysis

The primary aim of this study was to estimate trends in seropositivity rates of the adult Czech population. We estimated seroprevalence rates and 95% confidence intervals, we also standardized the seroprevalence rates by age and sex, using the Czech population as a standard. We used a multivariate Poisson regression model with a robust error variance to estimate the ratio of seroconversion and vaccination associated with sociodemographic indicators. Differences in prevalence were expressed as prevalence rate ratios (PRRs). We used standard descriptive statistics to characterize the study data set.

Population data on COVID-19 were obtained from the Czech Central Information System of Infectious Diseases (ISID), which includes records of all consecutive patients with COVID-19 in the Czech Republic identified and confirmed by laboratory testing. ISID data are routinely collected in compliance with Act No. 258/2000 Coll. on the Protection of Public Health and are publicly available in aggregated and anonymized form of open or authenticated data sets. All analyses were conducted using Stata version 15.1 (StataCorp, College Station, Texas 77845 USA).

## Results

This report is based on data from 22,130 subjects who participated in both phases of the study and therefore had repeated antibody measurements. Characteristics of the analytical sample are shown in **Table 1**. Just under 20% were under 40 years of age and 23% were older than 60 years, 62% were females and 43% of participants had tertiary educational level, and 65% (14,483) subjects reported vaccination by one of the four vaccines Comirnaty (BioNTech Manufacturing GmbH, Mainz, Germany), Spikevax (previously COVID-19 Vaccine Moderna; Moderna Biotech Spain, S.L., Madrid, Spain), Vaxzevria (previously COVID-19 Vaccine AstraZeneca; AstraZeneca AB, Södertälje, Sweeden), Jcovden (previously COVID-19 Vaccine Janssen; Janssen-Cilag International NV, Beerse, Belgium) available in the Czech Republic. The proportion of vaccinated persons increased with increasing age and increasing body mass index while it was lower in previously seropositive subjects. On the other hand, there was little variation in seroprevalence by sex and among ages groups. The proportion of self-reported vaccination was similar to official figures for the general population in the Czech Republic for September 2021 (see **Figure 1**).

**Table 1.** Characteristics of the study sample and proportions and prevalence rate ratios of seropositivity and vaccination.

|  | No. of participants | No. of seropositive | No. of vaccinated participants | Model of antibodies of any origin<br>(N = 22,130) |  |  | Model of propensity to vaccination<br>(N = 22,130) |  |  | Model of antibodies in unvaccinated participants<br>(N = 7,647) |  |  |  |
| --- | --- | --- | --- | --- | --- | --- | --- | --- | --- | --- | --- | --- | --- |
|  |  |  |  | % of seropositive | PRR (CI) | p value | % of vaccinated | PRR (CI) | p value | N of participants | % of seropositive | PRR (CI) | p value |
| **Sex:** |  |  |  |  |  |  |  |  |  |  |  |  |  |
| Female | 13,824 | 12,067 | 8,844 | 87.29% | **1.00** | - | 63.98% | **1.00** | - | 4,980 | 67.25% | **1.00** | - |
| Male | 8,306 | 7,282 | 5,639 | 87.67% | **0.99** | 0.012 | 67.89% | **1.05** | <0.001 | 2,667 | 65.47% | **0.95** | <0.001 |
| **Age groups:** |  |  |  |  |  |  |  |  |  |  |  |  |  |
| 18-29 | 1,491 | 1,202 | 770 | 80.62% | **1.00** | - | 51.64% | **1.00** | - | 721 | 61.72% | **1.00** | - |
| 30-39 | 2,774 | 2,275 | 1,534 | 82.01% | **1.02** | 0.215 | 55.30% | **1.03** | 0.420 | 1,240 | 61.05% | **0.97** | 0.338 |
| 40-49 | 6,700 | 5,725 | 4,177 | 85.45% | **1.01** | 0.194 | 62.34% | **1.17** | <0.001 | 2,523 | 64.05% | **0.97** | 0.226 |
| 50-59 | 6,049 | 5,405 | 4,061 | 89.35% | **1.03** | 0.003 | 67.14% | **1.23** | <0.001 | 1,988 | 70.32% | **1.04** | 0.170 |
| 60+ | 5,116 | 4,742 | 3,941 | 92.69% | **1.05** | <0.001 | 77.03% | **1.37** | <0.001 | 1,175 | 74.81% | **1.09** | 0.001 |
| **Education** |  |  |  |  |  |  |  |  |  |  |  |  |  |
| basic | 1,952 | 1,744 | 1,295 | 89.34% | **1.00** | - | 66.34% | **1.00** | - | 657 | 70.02% | **1.00** | - |
| medium | 8,024 | 7,119 | 5,348 | 88.72% | **1.00** | 0.972 | 66.65% | **1.02** | 0.275 | 2,676 | 69.21% | **1.02** | 0.337 |
| high | 7,544 | 6,689 | 5,223 | 88.67% | **1.00** | 0.890 | 69.23% | **1.08** | <0.001 | 2,321 | 65.75% | **1.02** | 0.394 |
| missing | 4,610 | 3,797 | 2,617 | 82.36% | **0.97** | 0.003 | 56.77% | **0.87** | <0.001 | 1,993 | 63.07% | **1.00** | 0.923 |
| **COVID in history** |  |  |  |  |  |  |  |  |  |  |  |  |  |
| Seronegative | 11,352 | 8,935 | 7,882 | 78.71% | **1.00** | - | 69.43% | **1.00** | - | 3,470 | 36.54% | **1.00** | - |
| Seropositive<br>- no symptoms | 5,597 | 5,374 | 3,458 | 96.02% | **1.28** | <0.001 | 61.78% | **0.75** | <0.001 | 2,139 | 90.04% | **3.45** | <0.001 |
| Seropositive<br>- with symptoms | 5,181 | 5,040 | 3,143 | 97.28% | **1.32** | <0.001 | 60.66% | **0.78** | <0.001 | 2,038 | 93.28% | **3.58** | <0.001 |
| **BMI** |  |  |  |  |  |  |  |  |  |  |  |  |  |
| <18.5 | 256 | 197 | 134 | 76.95% | **1.00** | - | 52.34% | **1.00** | - | 122 | 52.46% | **1.00** | - |
| 18.5-24.9 | 8,192 | 6,964 | 5,038 | 85.01% | **1.04** | 0.127 | 61.50% | **1.09** | 0.141 | 3,154 | 63.44% | **1.17** | 0.009 |
| 25-29.9 | 8,080 | 7,167 | 5,488 | 88.70% | **1.05** | 0.077 | 67.92% | **1.15** | 0.020 | 2,592 | 68.36% | **1.18** | 0.006 |
| 30+ | 4,802 | 4,369 | 3,312 | 90.98% | **1.06** | 0.046 | 68.97% | **1.16** | 0.013 | 1,490 | 74.30% | **1.20** | 0.003 |
| missing | 800 | 652 | 511 | 81.50% | **0.98** | 0.515 | 63.88% | **1.18** | 0.017 | 289 | 52.25% | **0.95** | 0.498 |
| **NCDs in history** |  |  |  |  |  |  |  |  |  |  |  |  |  |
| No | 13,888 | 11,958 | 8,688 | 86.10% | **1.00** | - | 62.56% | **1.00** | - | 5,200 | 65.23% | **1.00** | - |
| Yes | 7,152 | 6,500 | 5,161 | 90.88% | **1.00** | 0.818 | 72.16% | **1.06** | <0.001 | 1,991 | 71.97% | **1.00** | 0.813 |
| missing | 1,090 | 891 | 634 | 81.74% | **1.02** | 0.266 | 58.17% | **0.91** | 0.002 | 456 | 59.21% | **1.12** | 0.005 |
| **Vaccination** |  |  |  |  |  |  |  |  |  |  |  |  |  |
| Vaccination No | 7,647 | 5,095 | 0 | 66.63% | **1.00** | - | 0.00% |  |  |  |  |  |  |
| Vaccination Yes | 14,483 | 14,254 | 14,483 | 98.42% | **1.52** | <0.001 | 100.00% |  |  |  |  |  |  |
| **Total** | **22,130** | **19,349** | **14,483** |  |  |  |  |  |  | **7,647** |  |  |  |

**Figure 1.**
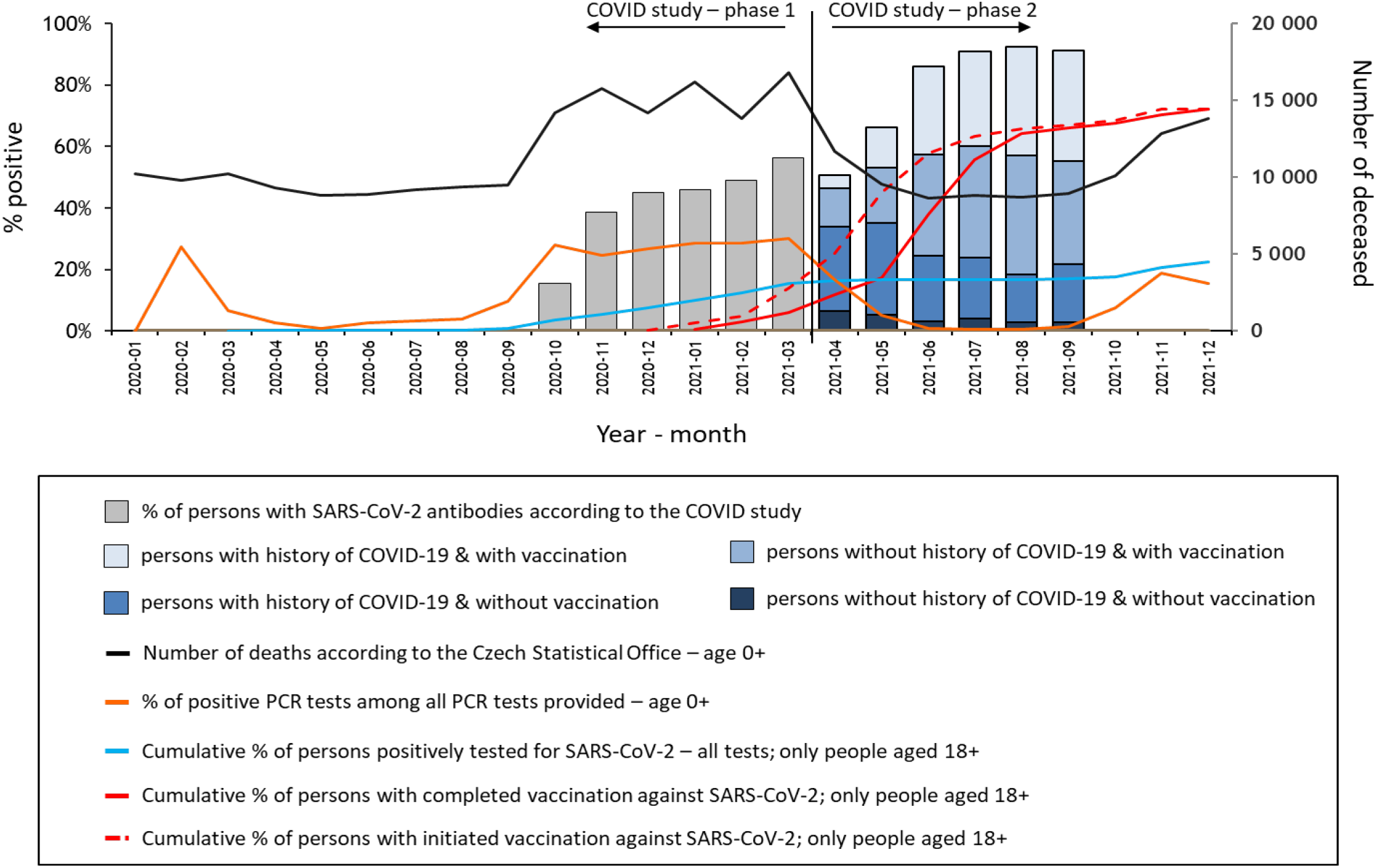
Temporal trends in indicators related to COVID-19 epidemic in the PROSECO Study and in the Czech national statistics.

Figure 1 shows the temporal trends in outcomes related to COVID-19 over both phases of the study. From March 2021 (end of phase 1), the seroprevalence increased from 56% to 91% in September 2021. While the rapid increase in seropositivity rates during phase 1 was due to natural infection, a substantial part of the increase during phase 2 was due to vaccination.

At phase one, 10,778 (49%) of participants were SARS-CoV-2 seropositive. Of the 11,352 seronegative subjects at phase 1, 1,009 reported positive PCR test between first and second blood sample (**Table 2**). **Table 3** shows seroprevalence rates at phase 2 by SARS-CoV-2 infection status at phase 1 and vaccination status. After standardisation to the Czech national population, the seroprevalence of anti-SARS-CoV-2 IgG antibodies was 24% among those who were seronegative at phase 1 and unvaccinated in phase 2; 90% among those who were seropositive at phase 1 or reported SARS-CoV-2 infection before phase 2; 97% among infection free before but vaccinated at phase 2, and almost 100% among those who both had SARS-CoV-2 infection before and were vaccinated at phase 2. In addition, only 9% of 4,367 unvaccinated subjects who were seropositive in phase 1 became seronegative over the 5-7 months until phase 2. From 7,495 SARS-CoV-2 immune naïve persons, only 210 (2.8 %) did not produce detectable IgG antibodies with 4-6 weeks after vaccination.

**Table 2.** Number of subjects with history of positive PCR test by seropositivity at Phase 1.

| Seropositivity at Phase 1 | SARS-CoV-2 infection reported (PCR) |  |  |  |
| --- | --- | --- | --- | --- |
|  | Prior 1<sup>st</sup> BS | Between 1<sup>st</sup> and 2<sup>nd</sup> BS | Never | Total |
| No | 1,080 | 1,009 | 9,263 | 11,352 |
| Yes | 6,397 | 95 | 4,286 | 10,778 |
| **Total** | **7,477** | **1,104** | **13,549** | **22,130** |

BS = blood sample collection

**Table 3.**
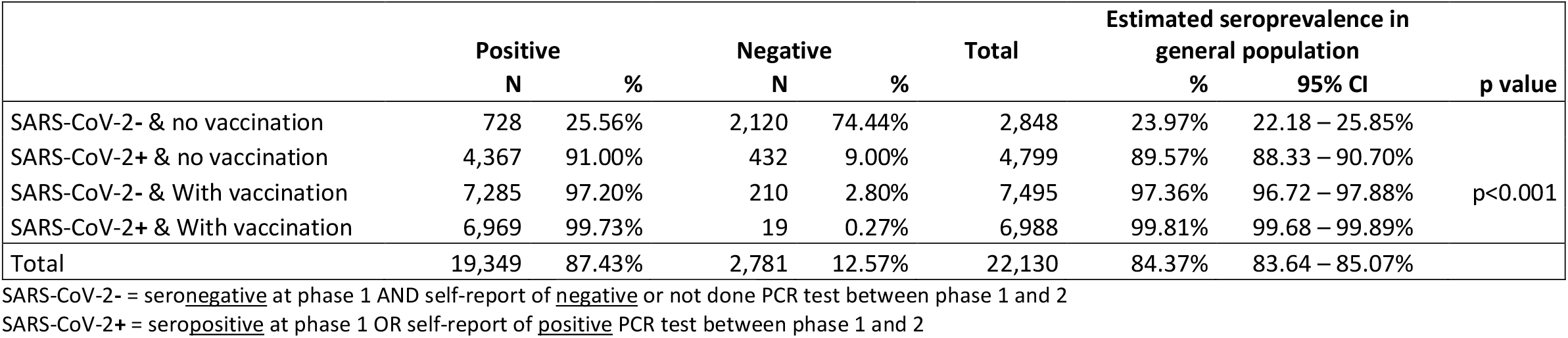
Seroprevalence at phase 2 by SARS-CoV-2 infection and vaccination status.

|  | Positive |  | Negative |  | Total | Estimated seroprevalence in general population |  | p value |
| --- | --- | --- | --- | --- | --- | --- | --- | --- |
|  | N | % | N | % |  | % | 95% CI |  |
| SARS-CoV-2- & no vaccination | 728 | 25.56% | 2,120 | 74.44% | 2,848 | 23.97% | 22.18 – 25.85% | p<0.001 |
| SARS-CoV-2+ & no vaccination | 4,367 | 91.00% | 432 | 9.00% | 4,799 | 89.57% | 88.33 – 90.70% |  |
| SARS-CoV-2- & With vaccination | 7,285 | 97.20% | 210 | 2.80% | 7,495 | 97.36% | 96.72 – 97.88% |  |
| SARS-CoV-2+ & With vaccination | 6,969 | 99.73% | 19 | 0.27% | 6,988 | 99.81% | 99.68 – 99.89% |  |
| **Total** | **19,349** | **87.43%** | **2,781** | **12.57%** | **22,130** | **84.37%** | **83.64 – 85.07%** |  |

SARS-CoV-2- = seronegative at phase 1 AND self-report of negative or not done PCR test between phase 1 and 2
SARS-CoV-2+ = seropositive at phase 1 OR self-report of positive PCR test between phase 1 and 2

## Discussion

In this prospective population-based study, we examined the changes in seroprevalence in a population-based sample with IgG antibodies measured twice, the second measurement being 5-7 month after the first on average. We found that after the rapid increase in seroprevalence during first phase (conducted in the 2^nd^ wave of the COVID-19 epidemic in the Czech Republic), there was further substantial increase in seroprevalence during the national vaccination campaign. By the end of phase 2 of the study, 91% of examined individuals had IgG antibodies against SARS-CoV-2; among vaccinated persons this proportion was over 97%.

### Strengths and limitations

The main methodological limitation of this study is the selection bias related to response rates. In phase 1, the response rates could not be established, since the number of persons who were invited by their insurance companies to participate in the study was known, as only the first 30,000 of those who attended were accepted in the study. These respondents were volunteers who were not entirely representative for the national population [8]. In addition, only about 74% of those who participated in phase 1 also participated in phase 2; as described in the methods, the phase 2 sample included slightly more women (62%) than the phase 1 had (61%).

Notwithstanding this limitation, the availability of repeated antibody measurements on a large number of individuals with high-quality chemiluminescent immunoassay is a major strength, since the prospective design allows assessment of antibody response in different groups of people. Both sex groups showed comparable seropositivity in both phases of the PROSECO study; the male and female rates in phase 1 (October 2020 to March 2021) were 46.1% vs. 47.2% due to natural infection, in phase 2 (April 2021 to September 2021) the rates increase to 87.7% vs. 87.3%, respectively, mostly due to vaccination.

Our results are in line with other national studies of antibody prevalence, such as the United Kingdom REACT-2 study[3], Blood donors study[6] and UK SARS-CoV-2 Immunity SIREN study[12]. In the week ending 28^th^ March 2021, which corresponds with the end of Phase 1 and the beginning of Phase 2 of the nationwide Czech PROSECO study, 55% of the adult population in England was tested positive for antibodies against the coronavirus SARS-CoV-2, these proportions were 49% in Wales, 59% in Scotland and 64% in Northern Ireland. The temporal trends were also comparable. By end of September 2021, the prevalence in England it was estimated as 92% of the adult population (and 90%, 91% and 91% in Wales, Scotland, and Northern Ireland, respectively (UK Office for National Statistics, www.ons.gov.uk). It is important to highlight that, unlike the Czech Republic, in the UK vaccination occurred earlier, before an increase in natural infection, resulting in less lost lives. By the end of Phase 2 in September 2021 seroprevalence increased to 91% in the Czech cohort.

Studies in other European countries have documented the built-up of seroprevalence in 2021, e.g., an 82% among German blood donors by September 2021 (Robert Koch Institut, SeBluCo-Studie). An Austrian cohort study of blood donors aged 18–70 years found that 10% of participants suffered with prior SARS-CoV-2 infection, and the seroprevalence of anti-SARS-CoV-2 IgG antibodies increased from 30% in March 2021 to 85% in September 2021 (n = 19,792), with the bulk of seropositivity due to vaccination. Anti-spike IgG seroprevalence was 99.6% among fully vaccinated individuals, 90% among unvaccinated individuals with prior infection and 12% among unvaccinated individuals without known prior infection[4 13]. Comparable results on blood donors were reported in the US, such as 20% for infection-induced antibodies and 83% for combined infection- and vaccine-induced antibodies in May 2021, and the estimated SARS-CoV-2 seroprevalence increased over time and varied by age, race and ethnicity, and geographic region[14].

Again, this is consistent with our findings. The highest seroprevalence in our study was seen among vaccinated persons with and without previous SARS-CoV-2 infection (99% and 97%, respectively), while the lowest seroprevalence was found among unvaccinated persons with no signs of disease. Moreover, only 2.8% of immune naïve persons did not produce detectable IgG antibodies with 4-6 weeks after vaccination. Furthermore, our prospective study also addressed the decline in antibody positivity after vaccination or after SARS-CoV-2 infection and we found that only among 9% of subjects who were seropositive in phase 1 became seronegative over the 5-7 months until phase 2.

In conclusion, the rapid increase in seropositivity during the 2^nd^ wave of the COVID-19 epidemic (covered by phase 1 of the PROSECO study) was followed by a similarly steep rise in seroprevalence during the national vaccination campaign, reaching seropositivity rates of over 97% among vaccinated persons in the Czech Republic in the period of April 2021 to September 2021. The combination of vaccination with the induction of a systemic immune response and natural infection with SARS-CoV-2 with the development of mucosal immunity is beneficial. It makes a significant contribution to good effect for diagnostic purposes and prophylaxis and leads to the development of protective immunity[15]. Seroconversion, as a marker of the ongoing immune response, is therefore an important measure of population immunity level to guide policy response and should play an important role in the WHO endorsed protocol for rapid adaptation and implementation of COVID-19 investigation[16].

## Data Availability

All data generated during the first and second phases of the PROSECO study is presented in this article. Anonymised data can be made available from the corresponding author upon request once all study phases are completed and data validated. Release of data is a subject of approval of the Ethical and Scientific boards of the PROSECO study.

## Acknowledgements

The PROSECO study was sponsored by the Prevention Programme of the Health Insurance Company of the Ministry of the Interior of the Czech Republic. The RECETOX Research Infrastructure was supported by the Ministry of Education, Youth and Sports of the Czech Republic (LM2018121), and VT and PP from the CETOCOEN PLUS project of ESIF (CZ.02.1.01/0.0/0.0/15_003/0000469). This work was supported from the European Union’s Horizon 2020 research and innovation programme under grant agreement No 857560 and the Ministry of Education, Youth and Sport of the Czech Republic/ESIF (CZ.02.1.01/0.0/0.0/17_043/0009632). This publication reflects only the author’s view and the European Commission is not responsible for any use that may be made of the information it contains. We thank all collaborating nurses, laboratories from the QualityLab association, and administrative personnel and especially the 30,054 participants who invested their time and provided samples and information for this study.

## Author contributions

VT, PP, LA and JK were responsible for the design of the study. KD, DK and LA were responsible for the study operation, coordination of data acquisition and quality management of participating laboratories. VT, PP and TP developed the operationalized research question and the statistical analyses plan. TP performed the statistical analyses. The first draft was written by VT and PP. MB contributed to the writing and finalizing of the manuscript. MB and HP provided expertise in epidemiology. All authors contributed to data interpretation, critically reviewed the first draft, approved the final version and agreed to be accountable for the work.

## Code availability

Statistical analyses were performed using STATA version 15.1 (StataCorp LLC, USA).

---

**Supplementary table S1:** Comparison of the persons participating in both phases with those who only attended phase 1.

|  | **Persons**<br>**participating in**<br>**both P1 and P2** | **%** | **Persons not**<br>**participating in P2** | **%** |
| --- | --- | --- | --- | --- |
| Total | **22,130** | 100% | **7,924** | 100% |
| **Sex** |  |  |  |  |
| Female | 13,824 | 62.5% | 4,438 | 56.0% |
| Male | 8,306 | 37.5% | 3,486 | 44.0% |
| **Age groups** |  |  |  |  |
| 18-29 | 1,491 | 6.7% | 1,069 | 13.5% |
| 30-39 | 2,774 | 12.5% | 1,485 | 18.7% |
| 40-49 | 6,700 | 30.3% | 2,431 | 30.7% |
| 50-59 | 6,049 | 27.3% | 1,658 | 20.9% |
| 60+ | 5,116 | 23.1% | 1,281 | 16.2% |
| **COVID in history** |  |  |  |  |
| Seronegative | 11,352 | 51.3% | 4,641 | 58.6% |
| Seropositive – no symptoms | 5,597 | 25.3% | 1,757 | 22.2% |
| Seropositive – with symptoms | 5,181 | 23.4% | 1,526 | 19.3% |
| **BMI** |  |  |  |  |
| under 18.5 | 256 | 1.2% | 85 | 1.1% |
| 18.5-24.9 | 8,192 | 37.0% | 2,791 | 35.2% |
| 25-29.9 | 8,080 | 36.5% | 2,360 | 29.8% |
| 30 and more | 4,802 | 21.7% | 1,189 | 15.0% |
| missing | 800 | 3.6% | 1,499 | 18.9% |
| **NCDs in history** |  |  |  |  |
| No | 13,888 | 62.8% | 6,563 | 82.8% |
| Yes | 7,152 | 32.3% | 539 | 6.8% |
| missing | 1,090 | 4.9% | 822 | 10.4% |

## References

1. Arora RK, Joseph A, Van Wyk J, et al. SeroTracker: a global SARS-CoV-2 seroprevalence dashboard. Lancet Infect Dis 2021;21(4):e75–e76 doi: 10.1016/s1473-3099(20)30631-9 [Published online First: 2020/08/09].

2. Vaughan A, Duffell EF, Friedl GS, et al. Seroprevalence of SARS-CoV-2 antibodies prior to the widespread introduction of vaccine programmes in the WHO European Region, January - December 2020: a systematic review. medRxiv 2021:2021.12.02.21266897 doi: 10.1101/2021.12.02.21266897.

3. Ward H, Whitaker M, Flower B, et al. Population antibody responses following COVID-19 vaccination in 212,102 individuals. Nature Communications 2022;13(1):907 doi: 10.1038/s41467-022-28527-x.

4. Siller A, Seekircher L, Wachter GA, et al. Seroprevalence, Waning and Correlates of Anti-SARS-CoV-2 IgG Antibodies in Tyrol, Austria: Large-Scale Study of 35,193 Blood Donors Conducted between June 2020 and September 2021. Viruses 2022;14(3) doi: 10.3390/v14030568 [Published online First: 20220309].

5. Stringhini S, Zaballa ME, Pullen N, et al. Seroprevalence of anti-SARS-CoV-2 antibodies 6 months into the vaccination campaign in Geneva, Switzerland, 1 June to 7 July 2021. Euro Surveill 2021;26(43) doi: 10.2807/1560-7917.es.2021.26.43.2100830.

6. Whitaker HJ, Elgohari S, Rowe C, et al. Impact of COVID-19 vaccination program on seroprevalence in blood donors in England, 2021. The Journal of infection 2021;83(2):237–79 doi: 10.1016/j.jinf.2021.04.037 [Published online First: 2021/05/11].

7. Soeorg H, Jõgi P, Naaber P, Ottas A, Toompere K, Lutsar I. Seroprevalence and levels of IgG antibodies after COVID-19 infection or vaccination. Infect Dis (Lond) 2022;54(1):63–71 doi: 10.1080/23744235.2021.1974540 [Published online First: 20210914].

8. Piler P, Thon V, Andrýsková L, et al. Nationwide increases in anti-SARS-CoV-2 IgG antibodies between October 2020 and March 2021 in the unvaccinated Czech population. Communications Medicine 2022;2(1):19 doi: 10.1038/s43856-022-00080-0.

9. Rostami A, Sepidarkish M, Leeflang MMG, et al. SARS-CoV-2 seroprevalence worldwide: a systematic review and meta-analysis. Clin Microbiol Infect 2021;27(3):331–40 doi: 10.1016/j.cmi.2020.10.020 [Published online First: 2020/11/25].

10. Bobrovitz N, Arora RK, Cao C, et al. Global seroprevalence of SARS-CoV-2 antibodies: A systematic review and meta-analysis. PLOS ONE 2021;16(6):e0252617 doi: 10.1371/journal.pone.0252617.

11. Grant R, Dub T, Andrianou X, et al. SARS-CoV-2 population-based seroprevalence studies in Europe: a scoping review. BMJ Open 2021;11(4):e045425 doi: 10.1136/bmjopen-2020-045425.

12. Hall V, Foulkes S, Insalata F, et al. Protection against SARS-CoV-2 after Covid-19 Vaccination and Previous Infection. The New England journal of medicine 2022;386(13):1207–20 doi: 10.1056/NEJMoa2118691 [Published online First: 2022/02/16].

13. Siller A, Wachter GA, Neururer S, et al. Prevalence of SARS-CoV-2 antibodies in healthy blood donors from the state of Tyrol, Austria, in summer 2020. Wien Klin Wochenschr 2021;133(23-24):1272–80 doi: 10.1007/s00508-021-01963-3 [Published online First: 20211026].

14. Jones JM, Stone M, Sulaeman H, et al. Estimated US Infection- and Vaccine-Induced SARS-CoV-2 Seroprevalence Based on Blood Donations, July 2020-May 2021. JAMA 2021;326(14):1400–09 doi: 10.1001/jama.2021.15161.

15. Russell MW, Moldoveanu Z, Ogra PL, Mestecky J. Mucosal Immunity in COVID-19: A Neglected but Critical Aspect of SARS-CoV-2 Infection. Front Immunol 2020;11:611337 doi: 10.3389/fimmu.2020.611337 [Published online First: 2020/12/18].

16. Bergeri I, Lewis HC, Subissi L, et al. Early epidemiological investigations: World Health Organization UNITY protocols provide a standardized and timely international investigation framework during the COVID-19 pandemic. Influenza Other Respir Viruses 2022;16(1):7–13 doi: 10.1111/irv.12915 [Published online First: 20211005].

